# Higher Maternal Education is Associated with Reduced Childhood Stunting in Zambia: Findings from the 2018 DHS

**DOI:** 10.1101/2025.06.10.25329376

**Authors:** Newton Nyirenda, Hannah Muturi, Caren Muyuni

## Abstract

**Background:** Childhood stunting remains a significant public health challenge in Zambia, contributing to poor cognitive development, increased morbidity, and long-term economic disadvantages. Maternal education is a known determinant of child nutrition outcomes, yet recent national-level data on this association are limited.

**Methods:** We analyzed data from the 2018 Zambia Demographic and Health Survey (DHS), focusing on children under five years of age. Stunting was defined as a height-for-age z-score below −2 standard deviations. Maternal education was grouped into four levels: no education, primary, secondary, and higher. Survey-weighted descriptive statistics and logistic regression models were used to assess the association between maternal education and child stunting, adjusting for sociodemographic and geographic covariates.

**Results:** Among 8,746 children with complete data, the overall prevalence of stunting was 34.3%. In adjusted models, children of mothers with higher education had significantly lower odds of being stunted compared to those whose mothers had no education (adjusted OR: 0.42, 95% CI: 0.27–0.66; *p* < 0.001). Primary and secondary education were not significantly associated with stunting after adjustment. Male sex, increasing child age, and lower household wealth were also significantly associated with higher odds of stunting.

**Conclusions:** Higher maternal education is independently associated with reduced risk of childhood stunting in Zambia. These findings underscore the importance of promoting women’s education beyond basic literacy as a long-term strategy to improve child health and nutrition outcomes.

## INTRODUCTION

Child undernutrition remains a major global public health challenge, with stunting defined as low height-for-age affecting approximately 22% of children under five years worldwide in 2022 (1). Stunting has long-term consequences including impaired cognitive development, reduced economic productivity, and increased risk of chronic diseases later in life (2,3). Sub-Saharan Africa bears a disproportionate burden, accounting for more than one-third of the world’s stunted children (1).

Maternal education is widely recognized as a critical determinant of child health and nutrition outcomes (4–6). Educated mothers are more likely to adopt health-promoting behaviors, utilize health services, and access and interpret nutritional information, thereby positively influencing their children’s growth (7–9). A previous multilevel analysis using 2013–14 Zambia DHS data found that higher maternal education was significantly associated with improved child nutritional status in Zambia (10). A more recent study using 2018 DHS data examined a broad range of determinants of stunting, including maternal education, but treated it as one of several contributing factors without focusing on its independent association (11). However, the strength and pattern of this association can vary by context and over time, and there is limited nationally representative evidence based on more recent data from Zambia.

Despite some progress, reductions in stunting in Zambia have not been consistent across all segments of the population. The prevalence declined from 40% in 2013–14 to 35% in 2018, but it remains among the highest in the Southern African region (12). Socioeconomic inequalities, including disparities in maternal education, may underlie these persistent gaps. Understanding how maternal education relates to stunting can inform targeted policy interventions aimed at vulnerable populations.

This study aimed to examine the association between maternal education and child stunting among children under five years in Zambia using data from the 2018 Demographic and Health Survey (DHS). We hypothesized that higher educational attainment among mothers would be linked to lower likelihood of stunting among their children, independent of other sociodemographic factors.

## METHODS

### Study Design

This analysis used cross-sectional data from the 2018 Zambia Demographic and Health Survey (ZDHS), a nationally representative survey conducted by the Zambia Statistics Agency in partnership with the Ministry of Health and the DHS Program. The ZDHS employed a two-stage, stratified cluster sampling strategy. In the first stage, enumeration areas were selected based on the 2010 national census sampling frame. In the second stage, a systematic sample of households was drawn from each cluster. Data collection involved structured interviews and anthropometric assessments of eligible children under five years of age (ZDHS 2018 final report).

### Study Population

The analysis included children aged 0–59 months who had valid anthropometric measurements and complete information on maternal education and relevant covariates. Children with missing or implausible height-for-age data (i.e., hw70 values of 9996, 9997, 9998, or 9999) were excluded from the analysis.

### Outcome Variable

The primary outcome was stunting, defined as height-for-age z-score (HAZ) below −2 standard deviations from the WHO Child Growth Standards. HAZ was derived from the DHS variable hw70, divided by 100 to scale values appropriately. A binary indicator variable was created for stunting (1 = stunted, 0 = not stunted).

### Exposure Variable

The main exposure was maternal education, categorized as: no education, primary, secondary, or higher. This variable was derived from DHS variable v106.

### Covariates

Covariates included child age in months (b19), child sex (b4), maternal age in years (v012), household wealth quintile (v190), urban or rural residence (v025), and geographic region (v101). Maternal body mass index (BMI), derived from v445 (and scaled by dividing by 100), was included in sensitivity analyses. All covariates were selected a priori based on their theoretical relevance to child nutrition and their potential role as confounders.

### Statistical Analysis

All analyses were conducted using R version 4.3.1 and incorporated the complex sampling design using the survey package. Sampling weights (v005), stratification variables (v022), and primary sampling units (v021) were applied to account for the survey design. From an initial sample of 9,959 children, 8,746 had complete data on stunting and key covariates and were included in the analysis, with 1,213 observations excluded due to missing values. The final analytic dataset was based on 545 clusters and 20 strata in line with DHS 2018 design specifications.

Descriptive summaries were generated using weighted means or proportions, depending on variable type. We fitted survey-weighted logistic regression models with a quasibinomial link to quantify the relationship between maternal education and child stunting, reporting both crude and adjusted odds ratios (AORs) along with their 95% confidence intervals. The adjusted model incorporated maternal education along with demographic and socioeconomic covariates. Children with incomplete data on any model variables were excluded from the regression analysis.

### Ethical Considerations

This study is based on a retrospective analysis of publicly available, de-identified data from the 2018 Zambia Demographic and Health Survey (ZDHS). The original ZDHS was conducted with ethical approval from the Tropical Diseases Research Centre (TDRC) in Zambia and the Research Ethics Review Board of the U.S. Centers for Disease Control and Prevention (CDC). Written informed consent was obtained from all adult participants during the survey. For participants aged 15–17 years, assent was obtained in addition to parental or guardian consent, in accordance with DHS protocols.

Access to the anonymized dataset was granted through the DHS Program. The data were obtained on 25 September 2024. At no point did the authors have access to information that could identify individual participants. As this analysis involved only secondary use of de-identified data, additional ethical approval was not required. Further details on DHS data protection and ethical procedures are available on their website.

## RESULTS

Maternal education is categorized as: No education, Primary, Secondary, and Higher.

Table 1 presents the characteristics of children under five and their mothers, stratified by maternal education level. Overall, the distribution of child sex was nearly equal across all education categories, though mothers with higher education had a slightly higher proportion of male children (55.6%) compared to those with no education (48.7%). A stark gradient was observed in place of residence: while only 16.1% of mothers with no education resided in urban areas, this figure rose progressively with education, reaching 81.7% among those with higher education.

**Table 1.** Characteristics of Children Under Five and Their Mothers by Maternal Education Level in Zambia, DHS 2018.

| Variable | Category | No Education (%) | Primary (%) | Secondary (%) | Higher (%) |
| --- | --- | --- | --- | --- | --- |
| **Child's Sex** | Female | 51.3 | 50.7 | 50.0 | 44.4 |
|  | Male | 48.7 | 49.3 | 50.0 | 55.6 |
| **Place of Residence** | Urban | 16.1 | 21.7 | 54.7 | 81.7 |
|  | Rural | 83.9 | 78.3 | 45.3 | 18.3 |
| **Child's Age (months)** | Mean | 29.5 | 28.4 | 27.6 | 28.6 |
| **Maternal Age (years)** | Mean | 31.8 | 29.2 | 26.6 | 31.5 |

\*Percentages are weighted and may not sum to exactly 100 due to rounding.

The average age of children ranged from 27.6 to 29.5 months across maternal education groups, with little variation. In contrast, maternal age showed more distinct differences: mothers with secondary education were younger on average (26.6 years), while those with no education or higher education were older (31.8 and 31.5 years, respectively).

Table 2 shows the unadjusted and adjusted odds ratios (ORs) for stunting among children under five, according to maternal education level. In the unadjusted model, children of mothers with secondary education had 25% lower odds of stunting compared to those whose mothers had no education (OR: 0.75; 95% CI: 0.61–0.93; *p* = 0.008). The odds were even lower for children of mothers with higher education (OR: 0.30; 95% CI: 0.20–0.45; *p* < 0.001).

**Table 2.** Association Between Maternal Education and Child Stunting in Zambia, DHS 2018.

| Maternal Education | Crude OR (95% CI) | p-value | Adjusted OR (95% CI) | p-value |
| --- | --- | --- | --- | --- |
| No education | 1.00 (ref) | — | 1.00 (ref) | — |
| Primary | 0.99 [0.82, 1.19] | 0.919 | 1.01 [0.83, 1.22] | 0.932 |
| Secondary | 0.75 [0.61, 0.93] | 0.008 | 0.84 [0.66, 1.08] | 0.174 |
| Higher | 0.30 [0.20, 0.45] | <0.001 | 0.42 [0.27, 0.66] | <0.001 |

After adjusting for child age, child sex, maternal age, household wealth, place of residence, and region, the protective association remained statistically significant only for higher education (adjusted OR: 0.42; 95% CI: 0.27–0.66; *p* < 0.001). The associations for primary and secondary education were attenuated and no longer statistically significant.

Table 3 displays results from the fully adjusted logistic regression model examining predictors of stunting among children under five in Zambia. Maternal education emerged as a significant protective factor: children whose mothers had attained higher education had 58% lower odds of being stunted compared to those whose mothers had no education (adjusted OR: 0.42; 95% CI: 0.27–0.66; *p* < 0.001). However, no statistically significant differences were noted for primary or secondary education levels.

**Table 3.** Fully Adjusted Odds Ratios for Factors Associated with Child Stunting, Zambia DHS 2018.

| **Variable** | **Adjusted OR (95% CI)** | **p-value** |
| --- | --- | --- |
| Reference | 0.74 [0.47, 1.15] | 0.175 |
| Primary | 1.01 [0.83, 1.22] | 0.932 |
| Secondary | 0.84 [0.66, 1.08] | 0.174 |
| Higher | 0.42 [0.27, 0.66] | <0.001 |
| Child age | 1.01 [1.00, 1.01] | <0.001 |
| Male sex | 1.41 [1.28, 1.56] | <0.001 |
| Maternal age | 0.99 [0.98, 1.00] | 0.071 |
| Poorer | 0.86 [0.74, 1.00] | 0.055 |
| Middle | 0.73 [0.61, 0.87] | <0.001 |
| Richer | 0.70 [0.56, 0.87] | 0.001 |
| Richest | 0.44 [0.33, 0.59] | <0.001 |
| Rural | 0.79 [0.66, 0.94] | 0.007 |
| Central | 0.92 [0.71, 1.20] | 0.464 |
| Copperbelt | 0.91 [0.74, 1.13] | 0.370 |
| Eastern | 1.40 [1.10, 1.78] | 0.006 |
| Luapula | 1.39 [1.11, 1.74] | 0.004 |
| Lusaka | 0.83 [0.62, 1.11] | 0.197 |
| Muchinga | 1.46 [1.16, 1.84] | <0.001 |
| Northern | 0.96 [0.68, 1.37] | 0.796 |
| North-Western | 0.84 [0.72, 1.10] | 0.148 |
| Western | 0.70 [0.55, 0.88] | 0.006 |

Age and sex of the child were also important determinants. Older children had slightly increased odds of stunting (adjusted OR: 1.01 per month; 95% CI: 1.00–1.01; *p* < 0.001), while male children were significantly more likely to be stunted than females (adjusted OR: 1.41; 95% CI: 1.28–1.56; *p* < 0.001).

Household wealth showed a clear inverse association with stunting. Compared to children from the poorest households, those in the richest quintile had less than half the odds of being stunted (adjusted OR: 0.44; 95% CI: 0.33–0.59; *p* < 0.001), with decreasing odds observed across successive wealth categories.

Unexpectedly, children residing in rural areas had slightly lower odds of stunting than those in urban areas (adjusted OR: 0.79; 95% CI: 0.66–0.94; *p* = 0.007). Geographic disparities were also evident: children in Eastern, Luapula, and Muchinga provinces faced significantly higher odds of stunting, while those in Western Province had reduced odds compared to the reference region.

## DISCUSSION

This study examined the association between maternal education and stunting among children under five in Zambia using nationally representative data from the 2018 DHS. We found that children whose mothers had higher education were significantly less likely to be stunted, even after adjusting for sociodemographic and household factors. Specifically, maternal attainment of secondary or higher education was associated with a substantial reduction in the odds of stunting, reinforcing the importance of maternal education as a determinant of child nutritional outcomes.

These results are consistent with earlier research conducted in Zambia and comparable low-and middle-income country settings. A multilevel analysis of the 2013–14 ZDHS also reported that higher maternal education was associated with improved child nutrition outcomes in Zambia (10). Similarly, a more recent study using the 2018 ZDHS found maternal education to be among the significant determinants of stunting, although it was analyzed in conjunction with several other risk factors such as birth weight, wealth, and breastfeeding practices (11). Unlike that study, our analysis focused specifically on maternal education and used a fully adjusted model to isolate its independent association with stunting.

The protective effect of maternal education may reflect multiple pathways. Educated mothers are more likely to have better knowledge of appropriate child feeding practices, improved hygiene behaviors, and greater utilization of antenatal and postnatal care services (13–15). They may also be better positioned to access and act on health-related information, manage household resources more effectively, and advocate for their children’s wellbeing in health and social systems (16,17).

In addition to maternal education, several other covariates emerged as significant predictors of stunting. Male children were more likely to be stunted than female children, consistent with patterns reported across sub-Saharan Africa (10,18). Child age was positively associated with stunting, likely reflecting the cumulative effects of nutritional deficiencies and infections over time. Wealth index was also a significant predictor, with children from wealthier households exhibiting markedly lower odds of stunting, underscoring the role of economic access to food and health services.

Regional differences in stunting also emerged. Children residing in Eastern, Luapula, and Muchinga provinces had significantly higher odds of being stunted compared to those in the Western province. These findings suggest persistent geographic inequalities in child health that may reflect regional disparities in food security, maternal education, and healthcare access (12,19). Addressing these disparities will require region-specific interventions that take into account local contexts and barriers.

Notably, while primary education alone did not significantly reduce the odds of stunting, higher education was strongly protective. This suggests that policies aimed at merely achieving basic literacy for women may not be sufficient to yield substantial improvements in child nutrition. Evidence from multi-country analyses indicates that at least ten years of maternal education, equivalent to completing secondary school, is often necessary to observe significant benefits for child growth and health outcomes(5). Expanding access to and completion of secondary and higher education among girls may offer more meaningful long-term health benefits for future generations.

This study has several strengths, including the use of a large, nationally representative sample and adjustment for a wide range of potential confounders. However, limitations include the cross-sectional design, which precludes causal inference, and the reliance on self-reported maternal education, which may be subject to recall or reporting bias. In addition, unmeasured factors such as maternal nutrition, birth weight, and household food insecurity could mediate or confound the observed associations.

## CONCLUSION

Addressing childhood stunting requires a multifaceted approach, and maternal education emerges as a key leverage point. This study found that children of mothers with higher education had significantly lower odds of being stunted, even after controlling for socioeconomic and demographic variables. While no association was found for primary education, secondary and higher education levels were associated with substantial reductions in stunting risk.

These findings reinforce the importance of promoting women’s education, especially at the secondary level or beyond, as a long-term strategy to improve child nutrition outcomes. Future efforts in Zambia should build on this evidence by strengthening education and nutrition policy linkages, ensuring that investments in girls’ education are recognized not only for their social benefits but also for their direct impact on child health.

## Data Availability

This study used publicly available data from the Zambia 2018 Demographic and Health Survey (DHS). The dataset is accessible from the DHS Program website (https://dhsprogram.com) upon request.

https://dhsprogram.com/data/dataset/Zambia_Standard-DHS_2018.cfm

